# Adaptive split ventilator system enables parallel ventilation, individual monitoring and ventilation pressures control for each lung simulators

**DOI:** 10.1101/2020.04.13.20064170

**Authors:** Shahroor Sarit Hadar, Sarouf Yarden, Oz-Ari Leav, M Gilad, K. Joseph, N Leshem, Z. Neta, M. Naor, B. Amir, L. Rotem

## Abstract

**Objective:** In mass crisis setting such as the COVID-19 pandemic, the number of patients requiring invasive ventilation may exceed the number of available ventilators. This challenge led to the concept of splitting ventilator between several patients, which aroused interest as well as a strong opposition from multiple professional societies (The joint statement)^1^.Establishment of a safe ventilator splitting setup which enables monitoring and control of each ventilated patient would be a desirable ability. Achieving independency between the Co-vent patients would enable effective coping with different individual clinical scenarios and broaden the pairing possibilities of patients connected to a single ventilator. We conducted an experiment to determine if our designed setup achieves these goals.

**Methods:** We utilized a double two limbed modified ventilator circuits which were connected to dual lung simulators. Adding readily available pressure sensors (transducers), PEEP valves, flow control valves, one-way (check) valves and HME filters made the circuit safe enough and suitable for our goals. We first examined a single lung simulator establishing the baseline set parameters, while monitoring ventilator measures as Tidal Volume. The initial ventilator setting we chose was a controlled mandatory ventilation mode with a PIP (peak inspiratory pressure) of 25cmH_2_O, PEEP (Positive End Expiratory Pressure) of 5 cmH_2_O. In pressure control set at 20 cmH_2_O, the recorded mean TV(tidal volume) was 1000 mL (approximately 500 mL/lung simulator) with an average MV(minute ventilation) of 13 L/min (or 6.5 L/min/lung simulator). After examining the system with the dual modified circuits attached, and obtaining all the ventilation parameters, we simulated several clinical scenarios. We simulated clinical events such as: partial or full obstruction, disconnection, air leak and compliance differentials, which occur frequently on a ventilation course. Thus, it is a paramount system demand to keep undisturbed ventilation to the Co-vent patient A, while being challenged by patient B.

**Results:** The adaptive split ventilator setup yields increased safety, monitoring, and controls ventilation parameters successfully for each connected simulated patient (using lung simulators).It also enables coping with several common clinical scenarios on a ventilation course, by allowing the care provider to control PIP and PEEP of each Co-Vent patient.

**Conclusion:** In a mass crisis setting, when there is a shortage of ventilators supply, and as a last resort, this setup can be a viable option to act upon. This experiment demonstrates the ability of the split ventilator to ventilate dual lung simulators with increased safety, monitoring and ventilation pressures control of each simulated patient. This split ventilator kept supporting a simulated patient with undisturbed parameters while the CO-vent patient was simulated to be disconnected, having an air leak, or exhibiting lung compliance deterioration. To the best of our knowledge, this is the first time a split ventilator setup demonstrates these capabilities. Our pilot experiment suggests a significant potential of expanding the ventilator support resources, and is especially relevant during COVID-19 outbreak. Since this setup has not been used in a clinical setting yet, further research should be conducted to explore the safety limits and the capabilities of this model.

## Introduction

In a mass crisis setting such as the COVID-19 pandemic, the number of patients who need invasive mechanical ventilation exceeds the supply of available units. A possible crisis standard of care strategy is the ventilation of two patients with a single mechanical ventilator. As pointed out by the “Joint statement”^1^, Co-venting strategy should only be considered as an absolute last resort^1^. The first descriptions of Co-venting multiple patients on a single ventilator were advanced by Neyman^2^ and Paladino^3^. The use of Co-Venting has been tested in controlled, experimental models using test lungs, animals or human volunteers for brief periods^4-12^. Lately New York State Department of Health approved the ventilator splitting to treat dual patients with one machine^13^. We conducted a study to determine if our setup can yield safe ventilation, monitoring, and control for each connected patient. We also challenged our setup with clinical scenarios that resemble common events on a ventilation course, assuring that no deleterious interactions between simulated patient’s circuits can occur. Our work consists of an experimental setup with pressure recording and a computational flow simulation model which describes the flow rates and different pressures throughout the system. The numerical model and pressure recordings enable a strong understanding of the system’s behavior in different scenarios which helps us shorten development times and approval processes.

## Methods

The experimental setup (**Figure 1**) consists of a single SERVO-s ventilator (Maquet, Solna, Sweden) double limbed (Hudson) tubing circuit which is connected to dual lung simulators. The lung simulators were used to simulate a single patient each, on the modified circuit in parallel operation. The ventilator circuit was modified by adding readily available components. Dual inspiratory inlet limbs (blue) and dual expiratory outlet limbs (red) were connected via standard Y splitters, plastic tubes and two lung simulators. The ventilator inspiratory and expiratory pressures as well as the lung simulator pressures were independently monitored and recorded by the data acquisition system. Each lung simulator can be connected to Flow sensor which can monitor the TV of each patient. Unfortunately we did not collect these measures in this experiment.Each inlet channel (colored blue in **Figure 1**) consists of:

**Figure 1:**
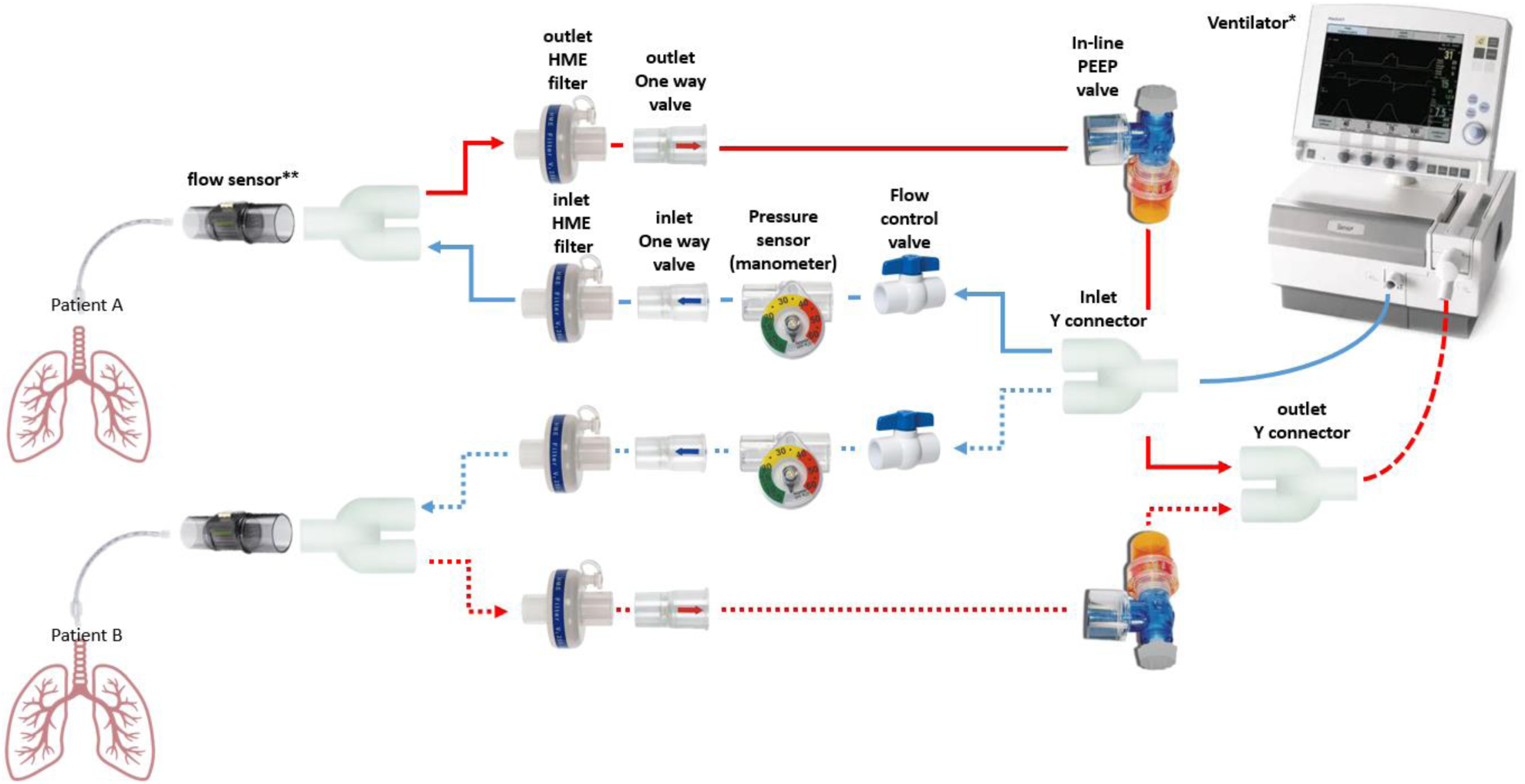
Scheme of the experimental setup.

- Flow control valve-used to set the PIP for each patient.
- Pressure sensor (manometer)-used to monitor the PIP and PEEP for each patient.
- Check valve-used to prevent backflow between the patients
- HME filter-for infection control purposes. Preventing cross contamination

Each outlet channel (colored red in **Figure 1**) consists of:

- In line PEEP valve-used to set and fine-tune the PEEP for each patient
- Check valve-used to prevent backflow between the patients, these valves are crucial for safety and independence of each circuit.
- HME filter-used to prevent cross contamination between patients

The setup is based on common, standard, readily available, medical grade components with small adjustments. A detailed list of all the essential components is in **Table 1**. The setup contains no added electronics or software, which makes it easy to understand and implement. The experiment configuration was a simulation of dual patients on a single ventilator in parallel operation (**Figure 1**).See a detailed setup photo in **Figure 2**

**Table 1:** list of essential components.

| Component | Quantity | Main technical properties |
| --- | --- | --- |
| Ventilator | 1 | MAQUET, servo-s* |
| Plastic tube | 3 m | 22 mm OD x 22 mm OD, <u>cuttable</u> cuffs every 6 in. |
| Y connector | 4 | 22 mm OD x 22 mm OD x 22 mm OD/15 mm ID |
| Flow control valve | 2 | 22 mm OD/15 mm ID x 22 mm OD/15 mm ID |
| One way valve | 4 | 22 mm ID Inlet x 22 mm OD Outlet |
| Manometer | 2 | 0 to 60 cmH<sub>2</sub>O |
| HME filter | 4 | 22 mm ID Inlet x 22 mm OD Outlet, |
| In-line PEEP valve | 2 | Adjustable, 0 to 20 cmH<sub>2</sub>O range |
| ** Proximal flow sensor | 2 | 22 mm ID Inlet x 22 mm OD Outlet, $\pm 250$ SLPM |

\* or similar
\*\* not demonstrated in our study

**Figure 2:**
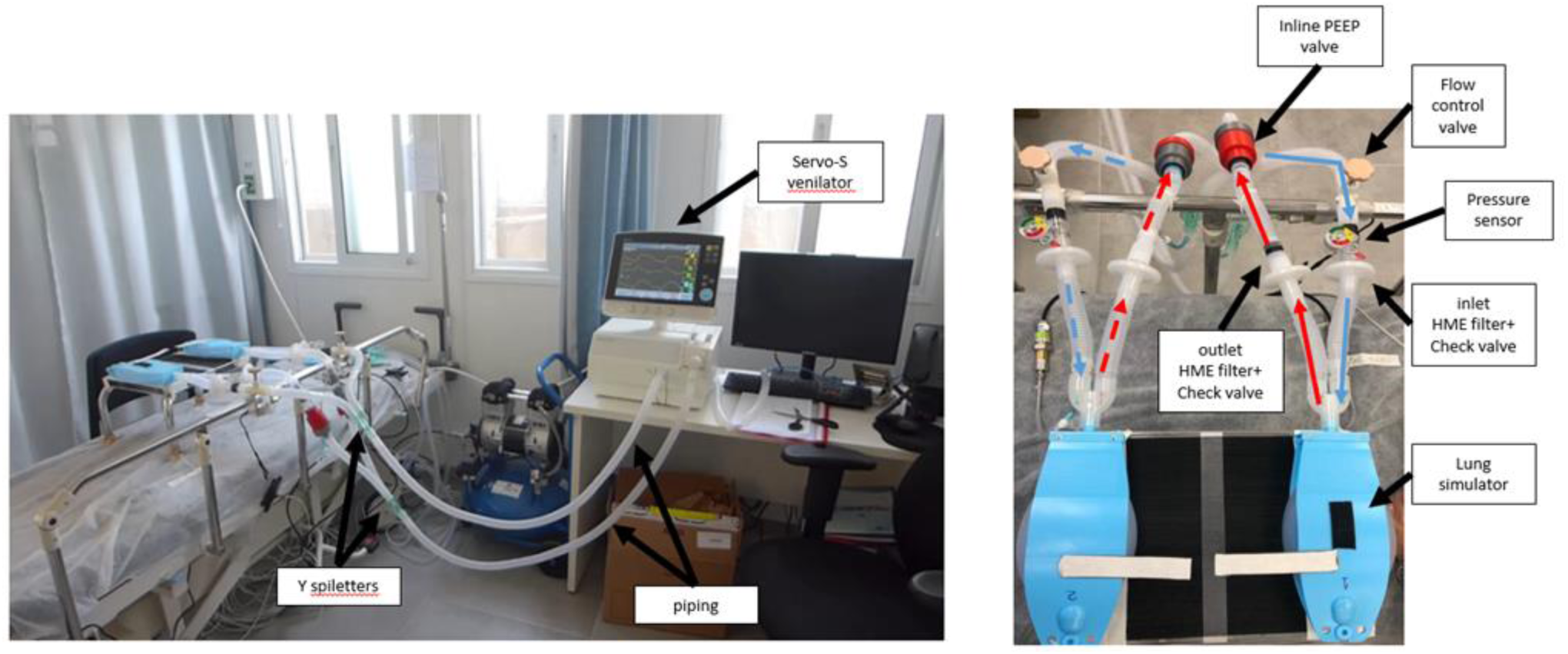
A detailed photo of the Adaptive split ventilator system.

### Ventilator settings, simulated clinical scenarios, and ventilation control

#### Ventilator settings

The initial ventilator settings were: controlled mandatory pressure ventilation (PC) with a PIP set at 25 cmH_2_O, respiratory rate (RR) of 16 breaths/min, PEEP of 5 cmH_2_O, FiO_2_ (Fraction of inspired oxygen) of 100%, I:E (Inspiratory to Expiratory ratio) for each breath 1:2. Using mandatory pressure-cycled ventilation can overcome most of the reservations raised by the Joint statement ^1^. This may allow splitting the ventilator in a reasonably safe manner, while limiting and control the driving pressure which suits a lung-protective strategy^7^. Using Volume Control mode for Co-venting provides no control over the TV or the maximal airway pressure of any patient. Thus on Volume Control ventilation, doubled TV may be delivered to an individual patient when his Co-Vent partner’s tube gets kinked, or obstructed. Patients sharing the ventilator on PC mode don’t have to match in compliance or by their size, as opposed to VC mode where the recommended TV is 6–8 mL/kg. The alarms setting were selected to mitigate risk to either patient and to identify any possible obstacles in the ventilation course right on occurrence. The sensitivity threshold was raised, to ensure no patient’s stimuli would occur, following our demand of no dependency between the patients.

First, we performed a system check with a single lung simulator, in order to identify leaks or inadequate settings, the set pressures provided a TV of 500ml/ lung simulator. In pressure control set at 20 cmH_2_O, the measured mean TV was 1000 mL (approximately 500 mL/lung simulator) with an average MV of 13 L/min (or 6.5 L/min/lung simulator). No more than 40 ml volume differential between the Inspiratory VT and Expiratory VT were noted. The ventilator display readouts were recorded. Simultaneously, the lungs simulators were subjectively inspected for symmetry of excursion. We also monitored objectively the height reached differences between the simulator lungs while being inflated –deflated on our ventilator. Specifically, we monitored for asymmetric inflation of individual lung simulators and incomplete deflation before subsequent inflation.

#### Simulated clinical scenarios

After examining the system with dual modified circuits connected to the ventilator and obtaining all the ventilation parameters, we simulated several clinical scenarios such as: partial and full obstruction, disconnection, air leak, and compliance differentials.

The simulations aim was to determine:

- Does the Adaptive Split Ventilator setup keeps ventilating the Co-vent patient B while patient A is having a clinical event?
- If so, does the Split Ventilator keep the same pre-event parameters?
- Does it alarm?
- Does it convey the right parameters and appropriate alarms for the event?

1. <u>Full obstruction</u> was simulated by placing a closing cover at the distal end of the circuit (at the supposed endotracheal tube connection). It also mocked a patient taking off the ventilator.
2. <u>Partial obstruction</u> was simulated by decreasing flow using the inserted flow valve in the same circuit.
3. <u>Simulation of air leak, disconnection of the circuit</u> (as self extubation) was resembled by opening the distal end of the circuit to room air.

#### Control of Ventilation parameters: PIP and PEEP

After simulating the clinical scenarios, we examined the ability to control the ventilator parameters as PIP and PEEP separately for each patient

- **PIP**: Simulation of PIP control as coping with deterioration of an individual patient. Sudden deterioration of patient A, who presents low compliance, could be overcome with raising PIP on the ventilator (to be delivered to patient A), while decreasing flow and subsequently the PIP pressure in patient B circuit, thus protecting patient B. By keeping the previous pressures for patient B, we are able to protect him, while patient A deteriorates.
- **PEEP** Simulation of differential PEEP control. Using the PEEP valve at one circuit enables us to increase PEEP of an individual patient while the Co-vent patient gets the previous PEEP of the ventilator.

## Results

### Ventilator settings

In this experiment we demonstrated that the Split Ventilator System could ventilate both lung simulators with equal excursions (subjective and objective measurements), with no leaks, no significant volume differentials, or backflow between the patients. The splitting ventilator recognized both lung simulators as single and revealed the TV sum of both, the appropriate Pressure-Volume curves, and flow curves. However, it is possible using the flow sensors (as in the setup scheme) to measure individual TV of each patient (not demonstrated in this experiment).

#### Simulated clinical scenarios

In the second part of the experiment we simulated common clinical scenarios, typical of a ventilation course. Ventilator alarms were carefully selected to mitigate risks to each patient, by setting alarms very close to parameters limits..

1. <u>Full obstruction/ patient A-clamped tube / disconnection off the ventilator.</u> (**picture A**) Simulated disconnection of patient A which may happen for a planned transport, weaning off the ventilator or suction procedure. In this simulation the split ventilator kept supporting patient B regardless of patient A circuit changes. Low MV alarms went off immediately, though patient B was ventilated with the same pressure parameters, as before.
2. <u>Partial obstruction of patient A tube with secretions/ kinked endotracheal tube</u> This event on a 1:1 patient ventilator support would cause TV decrease and MV alarms to go off. In the event of solid obstruction, as flow decreases, the patient would exhibit vital signs changes such as desaturation, tachycardia and End Tidal PCO_2_ reading decrement. As in single patient ventilation, the alarm of low MV went off, the inspiratory TV as the expiratory TV measures were lower than before. We demonstrated that the Split ventilator kept supporting patient B with undisturbed ventilation parameters. Setting the MV alarms value in tight gap from the mean MV ensured good control, and increased safety.
3. <u>Air leak (as self extubation) mocked by tube disconnection to room air (</u>**<u>picture</u> <u>B</u>**) On a 1:1 patient ventilator support the MV alarm would go off, Measured expiratory TV would be lower than the inspiratory TV, Vital signs alarms in the monitor would go off for desaturation and tachycardia. In the simulation, the split ventilator kept ventilating patient B with no interruptions or changes of his ventilator settings. A disconnection signal was ensured by setting the low MV alarm limits appropriately.

#### Control of Ventilation parameters: PIP and PEEP

After simulating the clinical scenarios, we examined the ability to control separately the ventilator parameters as PIP and PEEP for each patient

- **PIP**: In the simulation of patient A compliance deterioration we were able to raise PIP on the ventilator (delivered to patient A), while decreasing flow and subsequently the PIP in patient B circuit, thus protecting patient B (**picture C**). **Figure 4** presents the decreased flow and the subsequent lower reached PIP. The gradually increased pressure slope of patient B, reminds the gentle Pressure slope in PRVC (Pressure regulated volume control) mode, when the ventilator lengthens inspiratory rise time for achieving set pressure limits.
- **PEEP** By using the PEEP valve at one circuit, we were able to increase PEEP of an individual patient while the Co-vent patient got the PEEP set on the ventilator. The ability to control each PEEP while Co-venting, is a great measure for facing different PEEP needs (as in hypoxia, pulmonary edema, atelectasis, etc.) of the Co-venting patients

**Figure 3.**
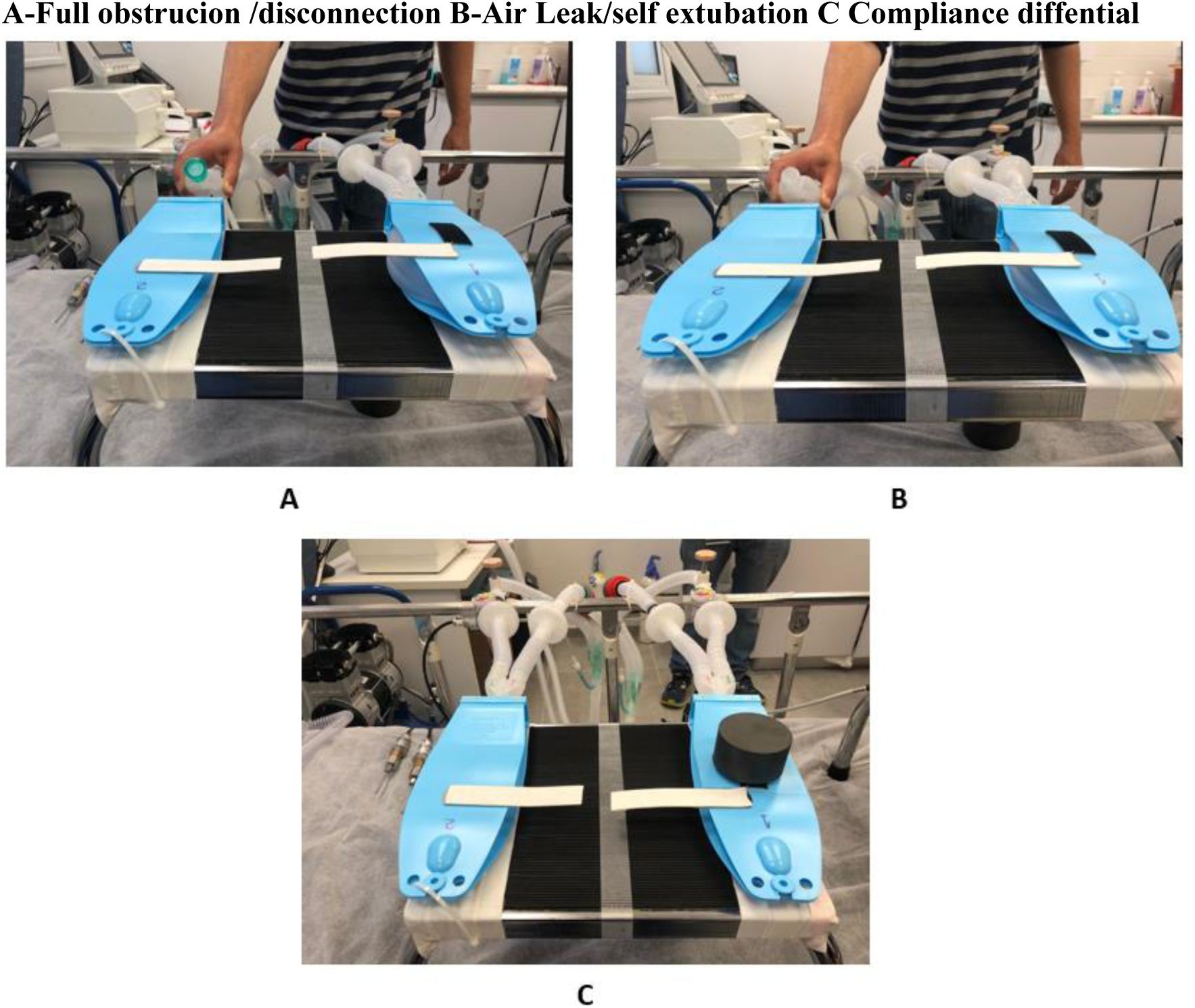
simulating common clinical events.

**Figure 4.**
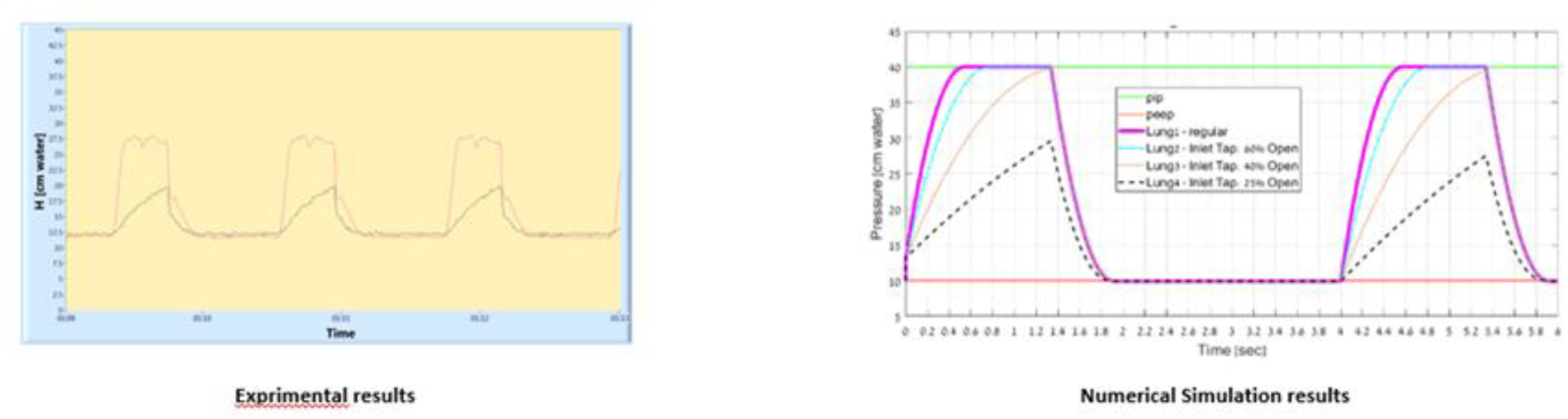
Experimental results of PIP control in simulation of compliance deterioration, compared to the expected results by computational flow simulation.

**Figure 5.**
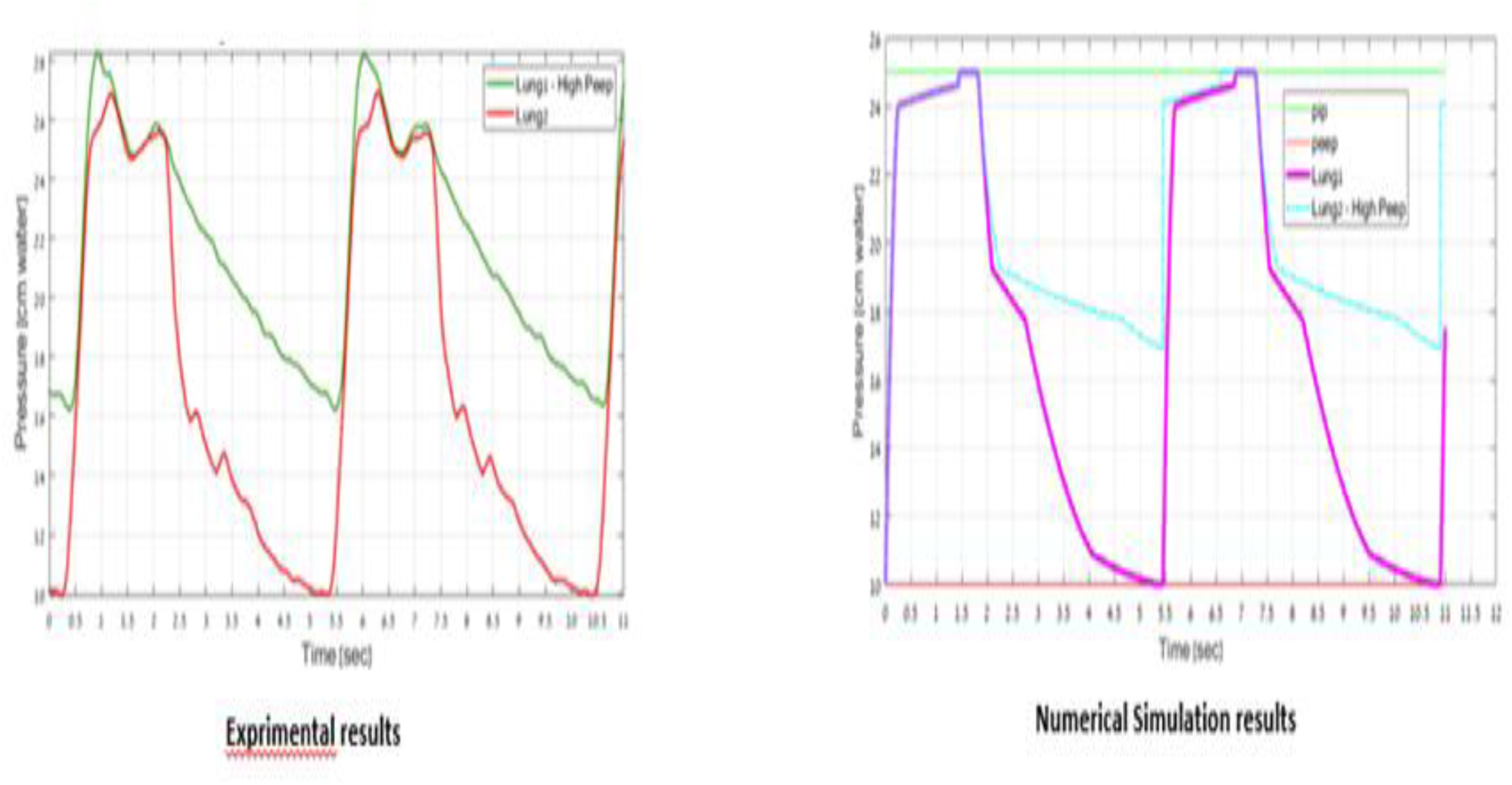
Experimental results of PEEP control compared to expected by the computational simulation results.

The ability to control each patient’s pressures as PIP and PEEP together with the advantages mentioned above, explore the extreme potential benefits of this system especially in dealing with a crisis such as COVID-19. This range of control abilities, safety, and monitoring would not only broaden the options to pair patients, but is also an exceptional tool to handle clinical differentials between paired patients.

## Discussion

In the COVID-19 Pandemic many hospitals will not be able to provide adequate supply of ventilators to all patients requiring invasive ventilation. Although the safe and reliable ventilation method is using one ventilator for one patient, in mass crisis setting Co-venting may be a viable option. Co-Venting has been tested only in controlled, experimental models using test lungs or animals for brief periods ^4^. Thus the use of Co-Venting should only be considered in a crisis situation. In this experiment we demonstrated that it is possible and considerably safer to simultaneously ventilate dual lung simulators, as simulated patients, using the split ventilator setup. This setup enables monitoring and ventilation parameters control of each simulated patient, with no possible deleterious interactions between simulated patient’s circuits. The relative independency of Co-venting patients is feasible due to the installed components of the circuit. The check one-way valves were crucial in this process, as the pressure transducers, the PEEP valves and the flow control valves. This experiment, successfully addresses the concerns of the “joint statement”.At the second part of our experiment we simulated several common clinical events on a ventilation course which challenge the used ventilator on a daily basis. The setup we manufactured proved to provide the safety and ventilating capabilities we were aiming for. The split ventilator succeeded in supporting each simulated patient in balanced ventilation distribution with no disturbances while being challenged with clinical events of Co-vent patient. The ventilation of the attached simulated patients is not affected by other individuals’ deterioration, obstructed tube, air leak, or disconnection. This setup was able to identify and manage the complexity of the Co-venting patients. We used a standard ventilator for installing this setup, combined with readily, available components with no electronics or software, so it is easy to understand, implement and operate.

The reliability and safety of Co-Venting in critically ill patients remains unknown. We believe that in this experiment the split ventilator setup addressed the challenging and the unpredictable aspects of Co-Venting. This is a significant step in developing a multi-patient ventilation system which may be a dominant, potentially lifesaving factor in facing the crisis of COVID -19.

## Limitations

The foremost limitation of this study is that it is a simulator study. Therefore, it could not stand for animal ventilation study with reassurance of blood gas tests. Still, the split ventilator exhibits reasonable safety, while monitoring and controlling each individual circuit parameters. It is also possible to measure each patient TV by using the flow sensors, already installed in our system, unfortunately we did not include these in this experiment. Despite the ability of our setup to monitor and control ventilator settings in differential lung compliances, it may not be achievable in every extreme situation. The infection control implications of Co-Venting are not firmly established. We used standard antimicrobial filters (HME) in each individual circuit limb. But further research is needed to fully evaluate the risk of cross contamination. The CDC states that the risk of Co-venting “with the above infection controlled interventions is likely to be small and would likely be appropriate in a crisis standard of care”. The split ventilator is safe only in Pressure Control Mode. Patients should not be able to trigger nor affect the ventilation of their Co-vent partners. We recommend increasing the sensitivity trigger threshold, sedating the patients and, as a last resort, paralyzing them. Auto-PEEP, increased dead space and resulted hypercapnea could be anticipated due to the increased length of the circuit. Since we can accept hypercapnea, as measures of lung protective strategy, we believe it should not be an obstacle for implementing our setup. All the Co-venting patients on the split ventilator would share parameters as RR, I: E, and FiO_2_.Co-venting patients on this setup could be considered only if all other alternatives are exhausted. Developing separate screen monitoring and control panel for each patient, would make this split ventilator even simpler and more user friendly operation. This setup is not compatible with every patient. Patients who suffer from baseline diseases as severe Asthma or COPD exacerbation should be precluded from Co-venting.

## Conclusion

In this experiment of the Adaptive split ventilator setup we were able to safely ventilate dual lung simulators while monitoring and having control of each simulated patient’s parameters. The split ventilator kept supporting a simulated patient with undisturbed parameters while the CO-vent patient was simulated to be disconnected, having an air leak, or exhibiting lung compliance deterioration. It also allows adjusting of PIP and PEEP for each Co-vent patient. To the best of our knowledge, this is the first time for a split ventilator to demonstrate these capabilities of increased safety, monitoring and ventilation parameters control for each simulated patient.

This Adaptive split ventilator setup has the potential to extend the ventilator support resources, especially during COVID-19 outbreak. Further research should be conducted on animals and humans in order to expand the safety and the capabilities of this model.

## Data Availability

https://www.sccm.org/getattachment/Disaster/Joint-Statement-on-Multiple-Patients-Per-Ventilato/Joint-Statement-Patients-Single-Ventilator.pdf?lang=en-US
https://www.gnyha.org/wp-content/uploads/2020/03/Ventilator-Sharing-Protocol-Dual-Patient-Ventilation-with-a-Single-Mechanical-Ventilator-for-Use-during-Critical-Ventilator-Shortages.pdf
https://medium.com/@pinsonhannah/a-better-way-of-connecting-multiple-patients-to-a-single-ventilator-fa9cf42679c6
https://www.hhs.gov/sites/default/files/optimizing-ventilator-use-during-covid19-pandemic.pdf

## Disclaimer

This Work (including all information contained herein), is provided for general informational purpose only with no warranty of any kind, either expressed or implied or statutory (including without limitations implied warranty of merchant ability, fitness for a particular purpose, receipt of any regulatory or other approvals, being within the standard relevant practices and non-infringement of third parties rights). The entire risk as to the above is on recipient and/or reader and/or user thereof (“Readers”) who shall be responsible to receipt all regulatory and other approvals required in relation with the use thereof and who shall be liable for all damages arising out of reliance, use, operation or disclosure thereof including any general, special, incidental or consequential damages. The receipt and/or use of the above, constitutes the consent to the terms set above

Readers are encouraged to confirm such information with other sources. The information was not approved by regulatory agencies

## Notes

### Competing Interest Statement

The authors have declared no competing interest.

### Funding Statement

The authors and their institutions did not receive and payment from a third party or services for aspect of the submitted work. There was not any external grants

